# Design and implementation of a system for automated monitoring of adherence to evidenced-based clinical guideline recommendations

**DOI:** 10.1101/2022.07.18.22277750

**Authors:** Gregor Lichtner, Claudia Spies, Carlo Jurth, Thomas Bienert, Anika Müller, Oliver Kumpf, Vanessa Piechotta, Nicole Skoetz, Monika Nothacker, Martin Boeker, Joerg J Meerpohl, Falk von Dincklage

## Abstract

**Background:** Clinical practice guidelines are systematically developed statements intended to optimize patient care. However, a gap-less implementation of guideline recommendations requires health care personnel not only to be aware of the recommendations and to support their content, but also to recognize every situation in which they are applicable. To not miss situations in which guideline recommendations should be applied, computerized clinical decision support could be given through a system that allows an automated monitoring of adherence to clinical guideline recommendation in individual patients.

**Objectives:** (1) To derive the requirements for a system that allows to monitor the adherence to evidence-based clinical guideline recommendations in individual patients, and based on these requirements, (2) to implement a software prototype that integrates clinical guideline recommendations with individual patient data and (3) to demonstrate the prototype’s utility on a COVID-19 intensive care treatment recommendation.

**Methods:** We performed a work process analysis with experienced intensive care clinicians to develop a conceptual model of how to support guideline adherence monitoring in clinical routine and identified which steps in the model could be supported electronically. We then identified the core requirements of a software system for supporting recommendation adherence monitoring in a consensus-based requirements analysis within loosely structured focus group work of key stakeholders (clinicians, guideline developers, health data engineers, software developers). Based on these requirements, we implemented a prototype and demonstrated its functionality by integrating clinical data with a treatment recommendation.

**Results:** Based on our conceptual flow chart model of recommendation adherence monitoring in clinical routine, we identified four main requirements of a software system for automated support of recommendation adherence monitoring of in-hospital patients: (i) Ability to interpret guideline recommendations’ semantics and logics, (ii) integration of clinical routine data from various underlying data structures, (iii) automatic adoption of new or updated guideline recommendations, and (iv) user interfaces optimized for distinct groups of users. Using a prototype implementation that fulfills these requirements, we demonstrate how such a system could be applied to monitor guideline recommendation adherence over time in clinical patients.

**Conclusions:** The four main requirements identified through our model-based analysis represent the most important aspects that need to be considered when developing a clinical decision support system for monitoring the adherence to evidence-based clinical guideline recommendations in individual patients. As each of the requirements corresponds to a different expertise (guideline development, health data engineering, software development, patient treatment), a modularized software architecture separated by area of required expertise seems favorable. Our prototype successfully demonstrates how such a modular architecture can be implemented to allow real-time monitoring of guideline recommendation adherence. This prototype, which we released as open source to invigorate collaboration, could serve as a basis for further development to integrate guideline recommendations with clinical information systems.

## Introduction

Clinical practice guideline recommendations are intended to optimize patient care by assisting decision making of health care professionals within specific clinical circumstances [1–3]. Thus, clinical practice guideline recommendations are among the most important potential clinical decision support tools [4,5]. Considering and implementing such recommendations during patient management is expected to be associated with improved patient outcome, especially in the case of evidence-based recommendations that were developed based on systematic reviews and appraisal of the available evidence [6–8]. However, a gap-less implementation of clinical practice guideline recommendations in daily routine work requires health care professionals not only to be aware of the existence of the respective guideline recommendations, to understand and support their content, but also to correctly recognize all situations in which specific recommendations should be applied [9].

Meeting the latter requirement becomes particularly demanding in the interdisciplinary treatment of patients with complex conditions that affect multiple organ systems, as it is often the case in critical care medicine [10–12]. To ensure that all health care professionals have an active knowledge about all guideline recommendations that apply in such situations and that they correctly recognize every situation in which these recommendations should be applied can prove difficult. Thus, treatment in critical care medicine is at a comparably high risk of deviating from guideline recommendations [10].

Besides the multitude of simultaneously applicable guideline recommendations in critical care, another aspect that can strongly affect guideline recommendation adherence is a high frequency of changes in recommendations [11]. The coronavirus disease 2019 (COVID-19) pandemic presented an exemplary situation in which dissemination and implementation of guidelines via conventional processes struggled to hold pace with the rapid development of recommendations and the speed at which recommendations were updated and changed over time [13,14].

To counter such difficulties and assist implementation of clinical guideline recommendations by means of computerized clinical decision support, various machine-readable guideline recommendation formalisms have been developed [15–22]. However, these formalisms focus on representing the finalized guideline recommendations and do not consider the systematic development process from which evidence-based guideline recommendations are derived. We have recently developed a *Fast Healthcare Interoperability Resources* (FHIR)-based formalism for the computer-interpretable representation of the whole guideline recommendation development process from developing systematic reviews of primary studies, rating the certainty of the available body of evidence, and finally applying evidence-to-decision frameworks to derive the final recommendation, called *Clinical Practice Guidelines on Evidence-Based Medicine on FHIR* (CPG-on-EBMonFHIR; [23]).

Based on this representation, the aim of this study was (1) to collect and analyze the requirements for providing clinical decision support via automated monitoring of individual evidence-based guideline recommendation adherence, and (2) to design & implement a prototype that fulfills the requirements and (3) to test the prototype’s applicability on real patient data.

## Methods

### Overview

To derive the requirements for a software system to monitor the adherence to clinical guideline recommendations, we first performed a work process analysis of the clinical processes that are to be supported by the system. After identifying and structuring these processes, we determined which subprocesses can be supported electronically and identified the requirements for how these subprocesses can be supported by a software system. Based on these requirements we designed a modular system architecture, which we implemented as an open-source prototype. To demonstrate the utility of the prototype implementation, we applied it to monitor the adherence to a COVID-19 treatment guideline recommendation based on clinical data from a large European university hospital.

### Work process analysis, identification of electronic support potential and requirements analysis

To derive the requirements for the system, we first conducted a needs analysis for the users of the system, the clinical staff. This needs analysis was conducted as a work process analysis, in which five experienced clinicians contributed to flow chart modeling of how the adherence to guideline recommendations would be monitored in clinical practice. Modeling was performed based on an iterative feedback process, in which for each iteration the model was adapted until no further changes were required.

Based on the work process model, we identified which parts of this process could be supported electronically and developed a flow chart model of how clinicians would interact with an electronic system to achieve the goal of monitoring patient-specific guideline recommendation applicability and adherence. Identification of support potential and process modeling was again performed in an iterative feedback process among the same group of clinicians together with a health software architect.

To identify the requirements of the software system for monitoring individual recommendation applicability and adherence, we performed a comprehensive needs analysis for the system by involving key stakeholders:

1. Clinical staff, as they are the primary users of the system.
2. Clinical practice guideline developers, as they create and maintain the guideline recommendations that are used by the system.
3. Health data engineers familiar with hospital IT infrastructure, as the system is required to process data from electronic health records.
4. Software developers, as they are required to build, test, and maintain the system.

Stakeholders were recruited by convenience from participants at senior level of their respective field within the COVID-19 evidence ecosystem (CEOsys) project of the federal network of university medical centers (NUM) in Germany [24]. They were individually approached and requested to participate. No compensation was offered for participation. We required at least two participants per stakeholder group.

### Software design and prototype implementation

The software prototype was implemented in an agile, rapid application development approach. The architecture followed a microservice pattern to allow efficient separation of concerns and scalable and exchangeable deployments of the system within the heterogeneous clinical IT infrastructures. Each container exposes a RESTful application programming interface (API) specified according to the OpenAPI 2.0 standard [25]. Backend modules were implemented in Python 3.8 and the frontend modules using RShiny [26].

### Demonstration of prototype utility

To demonstrate the utility of the prototype, we connected it to anonymized clinical data of a large university hospital (Charité - Universitätsmedizin Berlin, Germany) and integrated a recent strong evidence-based guideline recommendation for the treatment of patients with severe or critical COVID-19 [27]. The use of the anonymized clinical data for research was approved by the local ethics committee (Ethikausschuss 4 am Campus Benjamin Franklin, Charité—Universitätsmedizin Berlin, Chairperson Prof. R. Stahlmann, Application Number EA4/008/19, approval date: 06 Feb 2019, amendment date: 14 May 2020).

## Results

### Work process analysis

The needs analysis with the clinical staff resulted in a flow chart describing the work process of how the adherence to guideline recommendations should be monitored (Figure 2 a). The core insight of the work process analysis was that to evaluate and monitor guideline recommendation adherence, clinicians always work at the ward level and examine each patient individually to see whether a guideline recommendation applies and whether it is fulfilled.

**Figure 1:**
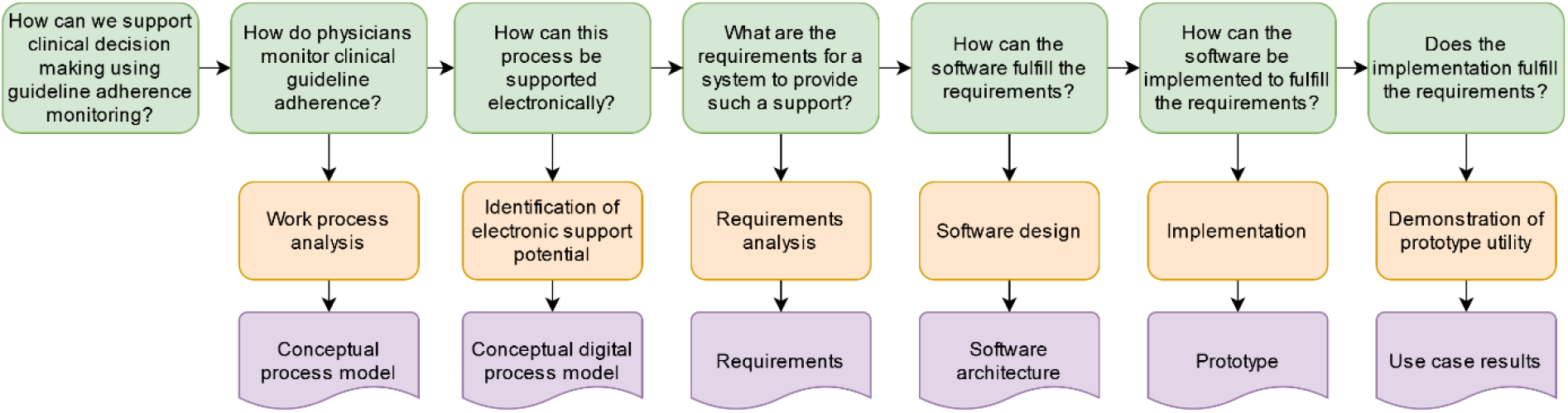
Flowchart of the study and the derived artefacts at each step.

**Figure 2:**
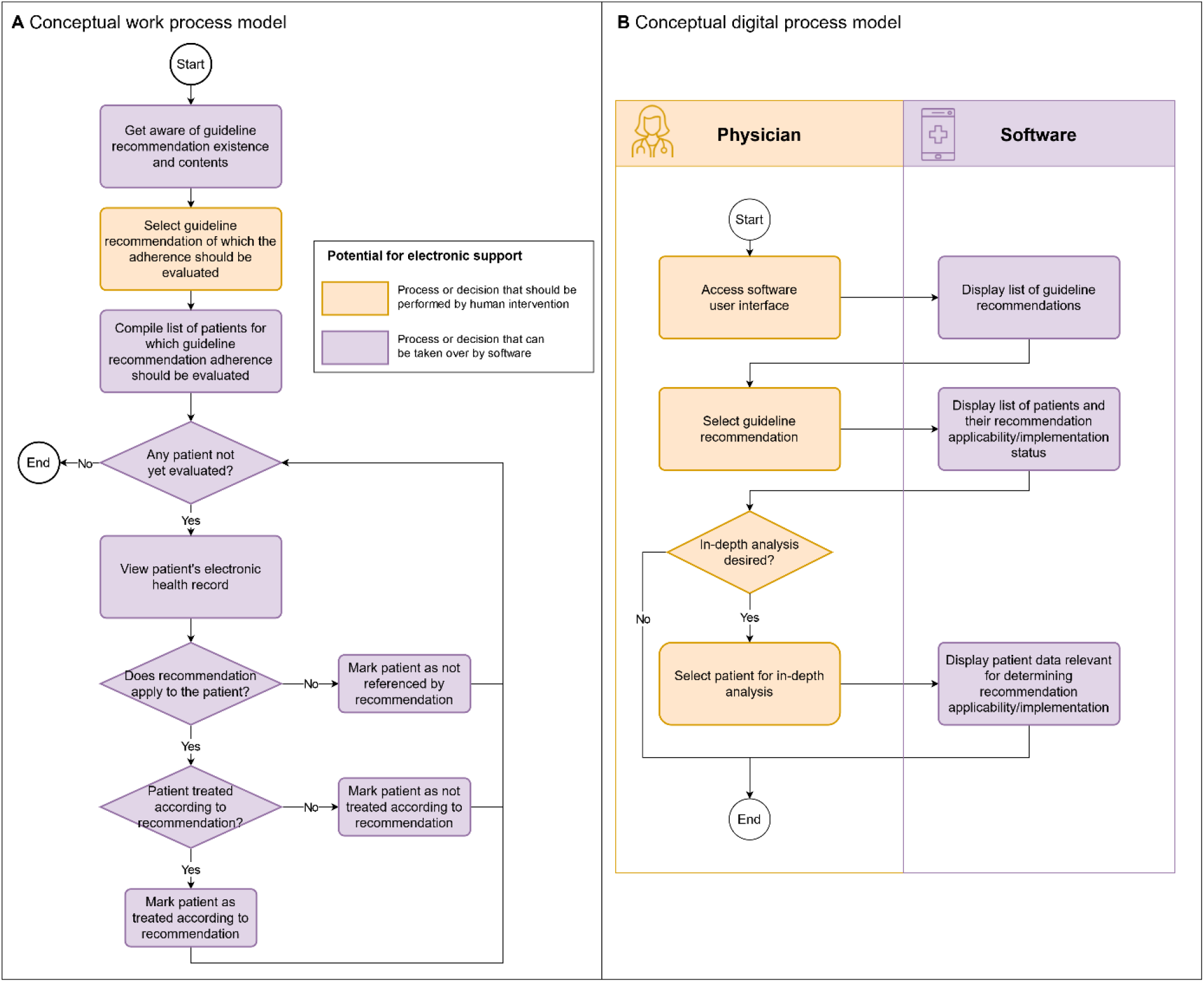
Conceptual process models of recommendation adherence monitoring in current clinical practice and using an electronic support system.

### Identification of electronic support potential

Based on the work process model, the clinical staff together with a health software architect identified which steps could be supported or covered by an electronic system. We identified nearly all steps as susceptible to being taken over by a software system (Figure 2 a). The same group then determined how this process should be supported electronically and developed a corresponding model of the digitized work process (Figure 2 b). The most important core insight of this analysis was the necessity to display the raw patient data that underlies the system’s decision on recommendation applicability/adherence, since clinicians using a software-based decision support system to monitor recommendation adherence want to be able to examine the raw patient data that underlies the system’s decision on recommendation applicability/adherence, to make sure that this data and decision is correct, as electronic health record data may contain errors.

### Requirements analysis

For a computer system that should support the defined work process (Figure 2 a), we identified four core requirements in a series of focus work group feedback rounds.

#### Requirement #1: The system needs to be able to decide whether a guideline recommendation is applicable and whether a guideline recommendation is implemented for a specific patient

The task of checking whether a guideline’s recommendation is applicable and whether it is implemented for an individual patient or not requires the system to be able to process both the semantical (i.e., what is the meaning of the words used in the recommendation) and logical (i.e., which of the words used in the recommendation define who the recommendation applies to and which words define what is to be done or not to be done) content of the recommendation. Therefore, the system needs to be provided with guideline recommendations in a format that is semantically correct, complete, and unambiguous.

We here focus on evidence-based guideline recommendations, which makes the decoding of the logical content of the recommendations particular easy: In the development process of evidence-based recommendations, it is standard practice to decompose the clinical question in consideration according to the PICO framework (population/patients, intervention, comparison, outcomes,) [28]. Therefore, in these recommendations, the patients to which the guideline recommendation is applicable to (P in PICO) and the intervention (I in PICO) that is recommended are distinctly defined at its best beginning with the systematic reviews supporting evidence-based guideline recommendations.

Regarding the decoding and interpretation of the semantical content of guideline recommendations, we have developed a FHIR-based format for the representation of clinical practice guideline recommendations to provide an interoperable, standards-based guideline recommendation exchange format that fulfills the above requirements [23]. Moreover, a variety of formalisms for representing guideline recommendations in a computer-interpretable way exist [29–31] and any of these could be used, provided that they are able to represent guideline recommendations semantically correct, complete and unambiguous.

#### Requirement #2: The system needs to be able to integrate clinical data from different data formats and data structures

Despite a multitude of initiatives for standardization, patient data is often only available in proprietary and non-standardized data formats and data structures that differ between countries, hospitals or even wards in the same hospital. Therefore, to make the system applicable across various existing IT infrastructural settings, the second core requirement is that it needs to accept data in a standardized, interoperable format, into which all proprietary data formats can be converted. Among data formats that fulfill these requirements are the OMOP common data model (CDM) [32] or FHIR-based formats (e.g. the US Core Profiles [33] or the German Corona Consensus dataset [34]).

#### Requirement #3: The system needs to automatically adopt changes in clinical guideline recommendations

Clinical guideline recommendations are subject to change as medical knowledge advances. Considering the vast number of new findings being published in the medical literature every day and the subsequent frequency of guideline recommendation updates, any efforts to manually implement updated guideline recommendations in a software system can be expected to delay updates and pose a source of errors [11].

One aspect that especially complicates error-free manual implementation of guideline recommendations in a software system is that such a task requires the expertise of at least two different and highly specialized fields: the expertise of the medical subspecialty providing the guideline recommendation and the software development expertise necessary for implementation into a system. Having both at one’s disposal for every single new or updated guideline recommendation is difficult and costly.

Therefore, the system needs to adopt changes in guideline recommendations without requiring changes in the system’s software code itself. Instead, once changes in guideline recommendation are released by the responsible medical societies or other appropriate sources, these changes should be automatically adopted by the system without the requirement of any manual changes in the system’s software.

#### Requirement #4: The system needs to provide user interfaces optimized for distinct user groups

Different users of the clinical decision support system require different user interfaces, depending on the specific work processes that are to be supported by the system. For example, medical or nursing staff working on individual patients require a system that is highly integrated with their standard patient data visualization used during the treatment process (e.g., the critical care information system used on the ward) to allow to integrate the decision support seamlessly into the individual patient care. In contrast, cross section staff like quality officers or supervising staff require more comprehensive overviews over multiple patients simultaneously, with less integration with other patient data, as their work processes that are to be supported by the system are more disconnected from individual patient care. Therefore, the fourth core requirement is that the user interface of the system must be customizable to meet the specific requirements that allow integration into the work processes of the respective groups of users that are to be supported.

### Prototype implementation

Considering the advantages of a modular system where each module corresponds to a specific specialty, we decided for a software architecture with four main modules that correspond to the previously described four main requirements that each require the involvement of only one of the three stakeholder groups besides the software developers (Figure 3; Table 1).

**Table 1:**
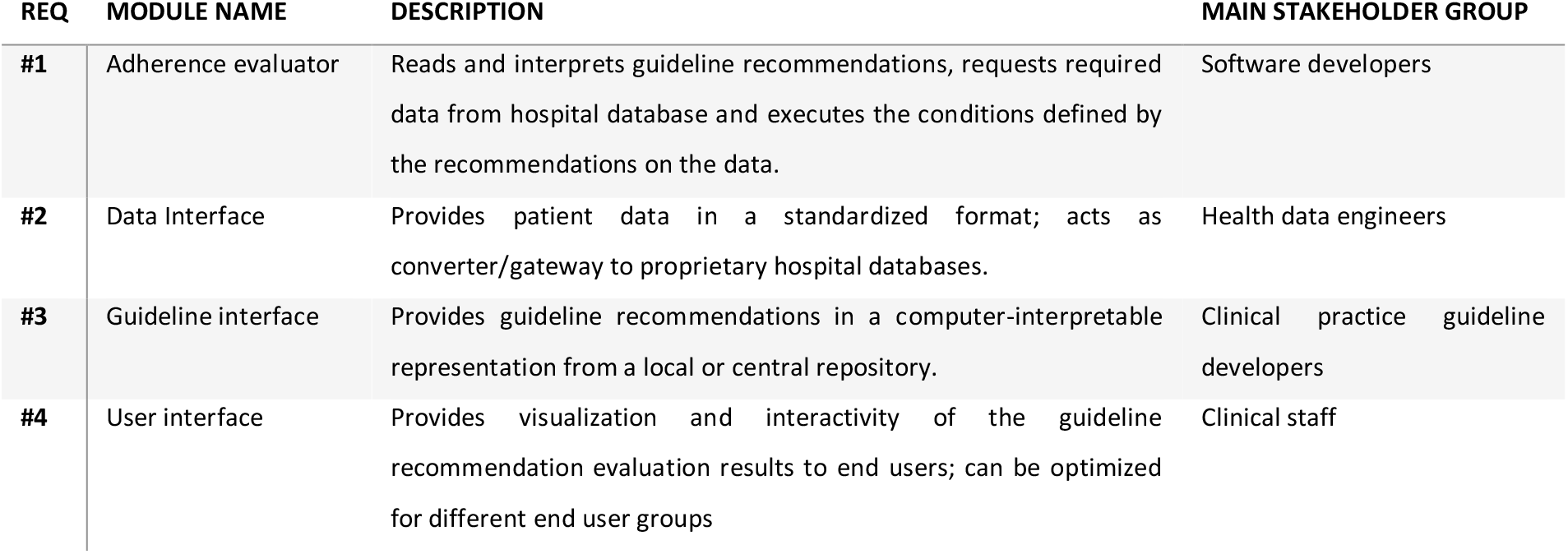
Overview of the prototype’s modules and their matching requirement.

| REQ | MODULE NAME | DESCRIPTION | MAIN STAKEHOLDER GROUP |
| --- | --- | --- | --- |
| #1 | Adherence evaluator | Reads and interprets guideline recommendations, requests required data from hospital database and executes the conditions defined by the recommendations on the data. | Software developers |
| #2 | Data Interface | Provides patient data in a standardized format; acts as converter/gateway to proprietary hospital databases. | Health data engineers |
| #3 | Guideline interface | Provides guideline recommendations in a computer-interpretable representation from a local or central repository. | Clinical practice guideline developers |
| #4 | User interface | Provides visualization and interactivity of the guideline recommendation evaluation results to end users; can be optimized for different end user groups | Clinical staff |

**Figure 3:**
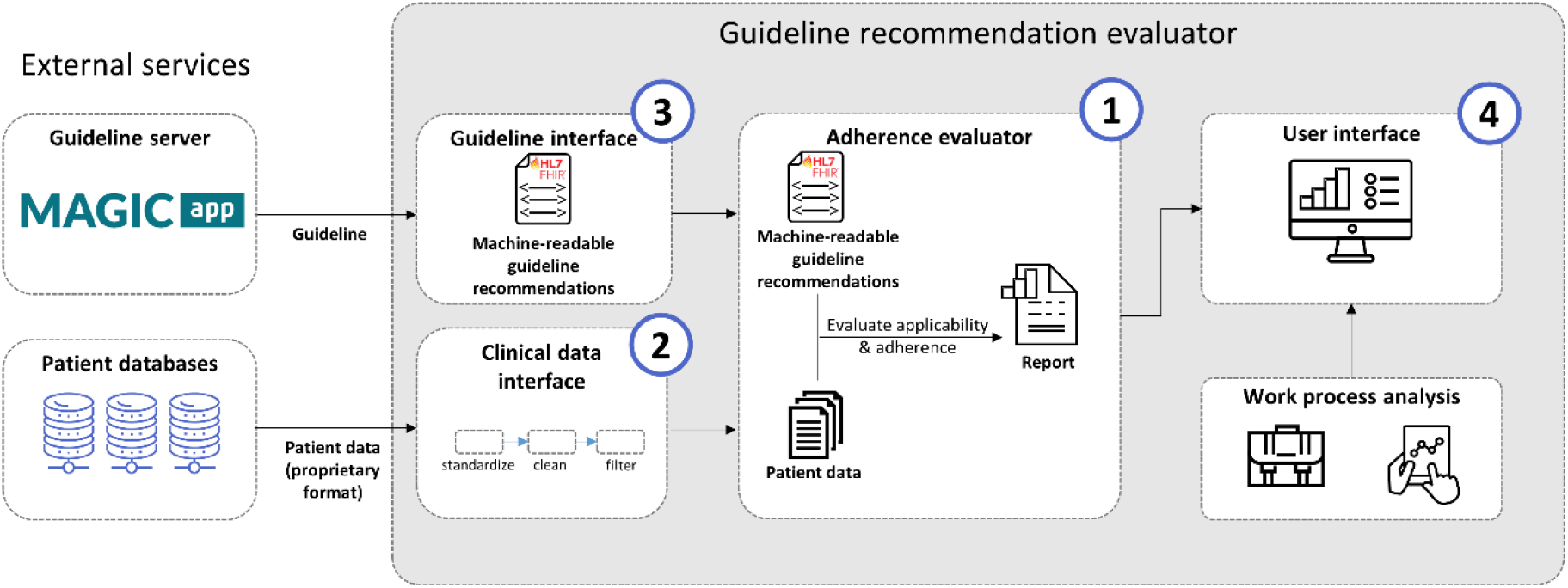
Architecture of the guideline recommendation evaluator for the automated integration of clinical guideline recommendations with real-time clinical data. Numbers indicate the requirement that is associated with the module.

The software was implemented as a containerized microservice architecture for effortless on-premises deployment within individual hospitals’ IT infrastructure, preventing any patient data from leaving the hospitals’ networks. We have designed the backend modules to be run within the same local network with the only outside access possible to the user interface backend. Thus, in our prototype implementation the user interface backend provides a user-level authentication to ensure only authorized users can access the output of the system. Depending on the actual deployment scenario, each of the microservices can be augmented by an authentication scheme.

For our exemplary prototype we have designed and implemented a user interface aimed to assist supervising medical staff in their task to review whether specific guideline recommendations are applicable and adhered to in individual patients that are currently treated in wards for which they are responsible (**Figure 4**). The user interface designed for this specific task allows the user to select the guideline recommendation to check and then, along with an overview over the patients currently treated in the ward, gives a condensed evaluation to which of the patients this specific recommendation is applicable and in which of the patients it is adhered to and in which it is not. Furthermore, the user interface allows to view the patient data on which the guideline recommendation evaluation was performed on to allow the clinician to review the evaluator’s results. The user interface is implemented as a dashboard website using RShiny [26], but is easily exchangeable by any other user interface framework or implementation due to the REST API interface of the user interface backend through which it receives data.

**Figure 4:**
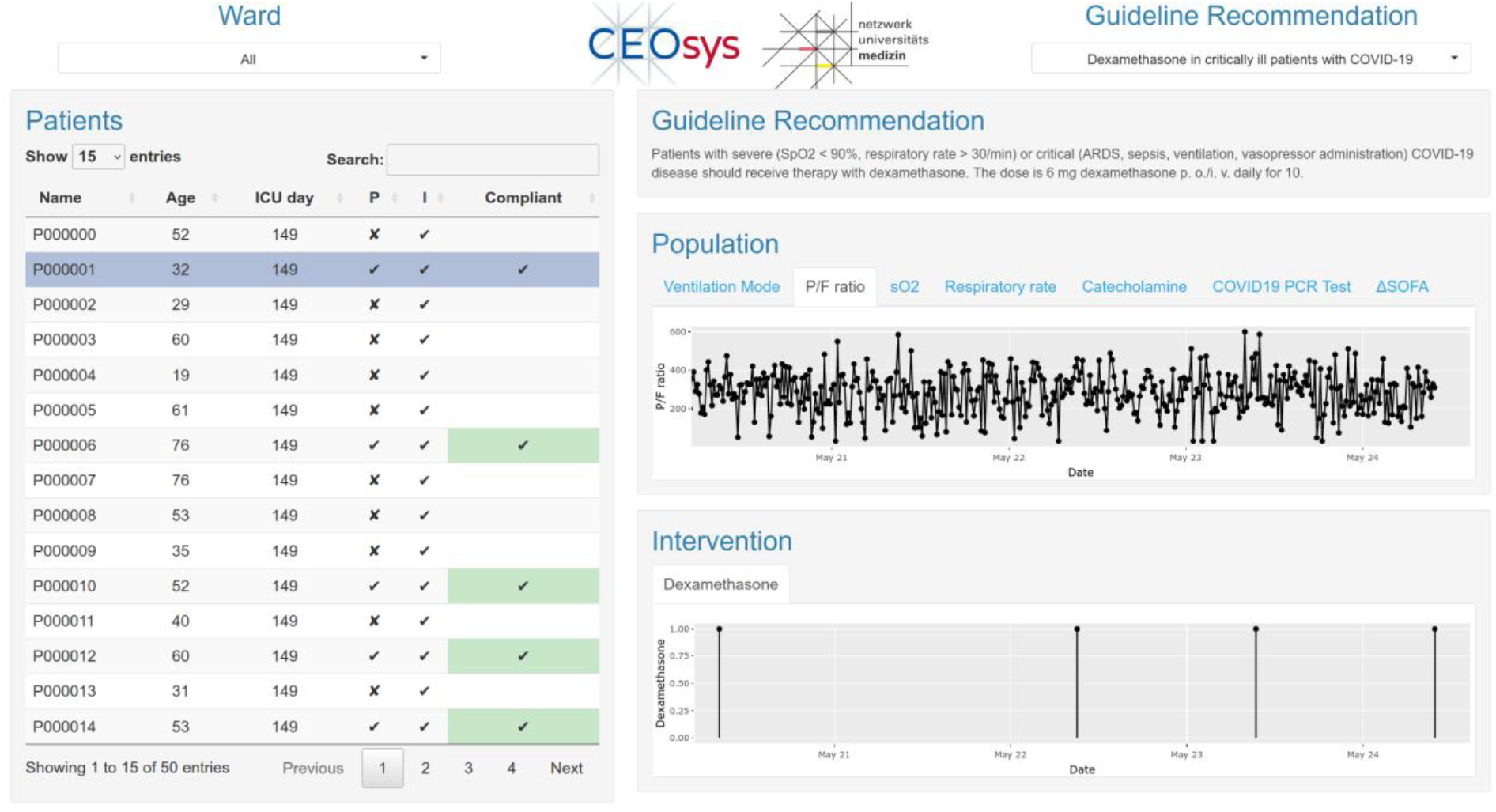
Prototype implementation of the user interface. The user can select a guideline recommendation of interest (top right) and view the patient-individual applicability and adherence of the recommendation on all current patients on a selected ward (left), where recommendation adherence is marked by check marks on green background. To allow the user to understand and review the results of the guideline recommendation evaluation, the user can select individual patients to show the original patient data required to assess the recommendation’s applicability and adherence (right).

### Demonstration of prototype utility

To demonstrate the utility of our prototype implementation, we have specified a recent strong evidence-based recommendation for the administration of dexamethasone to critically ill COVID-19 patients from the guideline for inpatient treatment of COVID-19 patients as a machine-readable guideline recommendation [35–37]. We connected the prototype to the critical care information systems and clinical information systems of Charité – Universitätsmedizin Berlin by means of a site-specific implementation of the clinical data interface. An exemplary time-dependent analysis of a guideline recommendation’s applicability and adherence is shown in Figure 5.

**Figure 5:**
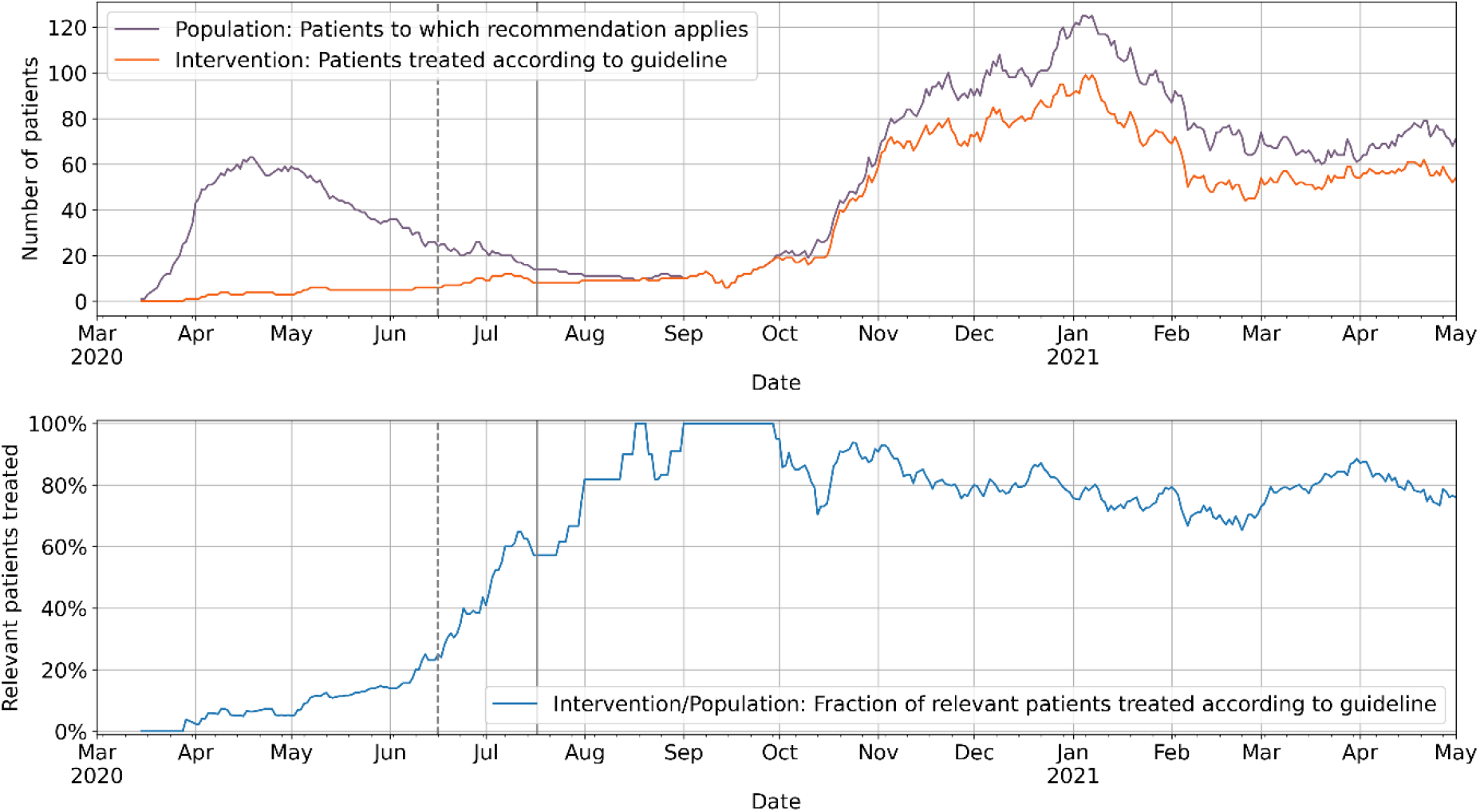
Individual applicability and adherence of a guideline recommendation to treat severe and critical COVID-19 patients with steroids. Top: Shown are the number of patients to which the guideline recommendation is applicable (purple) and which have been treated according to the guideline recommendation (orange) between March 2020 and May 2021. Bottom: Shown are the number of patients that are treated according to the guideline recommendation as a fraction of patients to which the guideline recommendation is applicable. Vertical gray lines indicate date of the first press release of the detected Dexamethasone efficacy in the RECOVERY trial (dashed line [38]) and the initial publication of the trial results (solid line; [39]).

## Discussion

In this paper, we demonstrated the system architecture and prototype implementation of a clinical decision support system that automatically integrates clinical guideline recommendations with real-time clinical data to assist health care professionals by visualizing whether guideline recommendations apply to individual patients and whether the guidelines recommendations have been followed in individual patients or not. We described our stepwise approach for the development of the system, including the core requirements which shaped our software architectural design, as well as our prototype implementation and demonstrated the prototype’s utility using a COVID-19 treatment guideline recommendation on clinical data.

To evaluate our architecture and prototype, we implemented a recent clinical guideline recommendation on treatment of patients with severe or critical COVID-19 disease and integrated the recommendation with data from a large university hospital to analyze the guideline recommendation adherence over time (Figure 5). The integration of the machine-readable guideline recommendation with clinical data could accurately detect the first and second wave of COVID-19 intensive care treatments [40] and the successful implementation of the guideline recommendation for the second wave, as seen by a >70% of relevant patients treated according to the recommendation. The non-100% guideline recommendation implementation in our specific data set is primarily due to the fact that a large part of COVID-19 patients treated in this university medicine center were transferred from other hospitals and in parts have been treated with steroids in accordance with the guideline recommendation before arriving on the intensive care unit of our hospital [41]. Such a situation in which the recommended treatment has already taken place but was recorded in a different patient data management system could be solved by increased interoperable data exchange between different health care providers.

An automated integration of guideline recommendations with clinical data as done by our developed systems holds several advantages: The system can provide a certain kind of decision support during individual patient treatment by pointing to applicable guideline recommendations, which the treating health care professionals might either not be aware of or which are known but whose applicability might go unnoticed. Additionally, the monitoring of guideline recommendation adherence across groups of patients provides possibilities for their use as quality and performance indicators [42,43] that can easily be monitored in real-time using a system as we propose it here. Another similar aspect could be the application of the system to monitor process implementation of new guidelines and to provide clinical insights in applicability of guideline recommendations that are useful for guideline updates. Independent from official clinical guidelines, the system can also easily be used to monitor hospital-specific treatment standards by formulating them as machine-readable guideline recommendations and providing them via the guideline interface to the adherence evaluator.

We have designed our system to require machine-readable guideline recommendations, however, guideline recommendations are currently available only in narrative, human-readable format. Thus, individual guideline recommendations need to be converted in a machine-readable format first, adding an extra amount of work. However, the specification of guideline recommendations in a machine-readable format enforces a precise and accurate formulation of guideline recommendation preventing ambiguities, as these could not (easily) be resolved or understood by a software system. Additionally, in contrast to converting human-readable recommendations into machine-readable recommendations, the generation of precise human-readable formulations of a guideline recommendation that is formulated in a machine-readable standard is a comparably simple task. Therefore, specifying guideline recommendations from the start in a machine-readable format holds multiple advantages and we therefore consider it a desirable change in the current practice of high quality, evidence-based guideline recommendation development. An alternative approach to using a machine-readable guideline recommendation specification could be the application of recent advances of natural language processing (NLP) methods to allow the computer to “understand” and process human-readable guideline recommendations [44,45]. However, any errors unknowingly introduced by such an approach (e.g., due to imperfect “understanding” of the guideline recommendation by the NLP algorithms) could have severe consequences on patient health and outcomes. Therefore, we believe that the explicit statement of guideline recommendations in a standardized machine-readable format is currently the more suitable choice.

In our prototype implementation we have used an EBMonFHIR-based representation of clinical guideline recommendations that covers the whole development process of evidence-based guidelines, from the underlying systematic review of available evidence over the rating of the individual evidence to the final recommendation in a computer-interpretable way [23]. The computer-interpretable links to the development process are particularly useful in meeting the requirement of the system to adapt to updated guideline recommendations (requirement #3), as updates to recommendations based on new evidence or re-appraisal of existing evidence can be automatically passed to the system without the need of newly and manually converting human-readable guideline recommendations into a representation formalism. While requirement #3 could in principle be met by any guideline recommendation formalisms that is semantically correct, complete, and unambiguous, the CPG-on-EBMonFHIR representation offers an advantage especially with guideline recommendations that are updated regularly, such as recommendations from living guidelines. Additionally, the CPG-on-EBMonFHIR representation could allow the users of the software system to evaluate the certainty of the evidence and the evidence-to-decision process underlying individual recommendations. We did not include this functionality in the prototype implementation, which, however, may become part of a later extension stage of the system implementation.

In our approach, the user interface is separated from the rest of the software to allow user- and context-specific implementation of user interfaces. We have provided a prototype as a dashboard website to demonstrate its feasibility. However, in clinical practice during individual treatment it might be desirable to integrate the suggestions of guideline recommendations into the clinical information system (CIS) that is implemented on the ward. Due to the separation of user interface and backend in our system, these integrations could be readily implemented depending on the CIS, e.g. as SMART-on-FHIR clinical decision support (CDS) hooks [46,47], if the CIS provides a SMART-on-FHIR interface.

We have designed our software system to retrieve guideline recommendations from a centralized repository, which could be hosted by medical societies or national standardization organizations. This allows these societies or organizations to develop guideline recommendations independently of our system, thereby focusing on their expertise. Once new or updated guideline recommendations are published by the medical societies or organizations on their servers, they can be automatically retrieved by our system and integrated with clinical data. However, a checkpoint in this process should be established, where health care professionals of the individual hospitals first review new or updated guideline recommendations retrieved from the central repository before releasing them for implementation in their hospital. This helps to mitigate risks that arise if the central guideline recommendation server is compromised by malicious attackers and also ensures that the guideline recommendations implemented in the hospitals are in accordance with the hospitals’ policies.

## Conclusion

In this work, we have designed a system that integrates guideline recommendations with real-time clinical data to evaluate individual guideline recommendation adherence and developed a functional prototype. The proposed system holds advantages both for the individual treatment of patients, as clinical guidelines condense the current state of medical knowledge into treatment recommendations, and for quality management and monitoring of patient treatment standards in hospitals. We have specified a modular software architecture where each module corresponds to a particular area of expertise, allowing experts from different fields (guideline developers, software engineers, medical data engineers and health care professionals) to work independently and focus on their area of expertise. We have released the source code of our system under an open source license (see code availability) and invite for cooperation and collaborative further development of the system [48].

## Data Availability

All code produced is available online at https://github.com/CEOsys/grevaluator.

## Code availability

The complete code of the clinical guideline recommendation system is published under an open-source license (AGPL v3.0) at https://github.com/CEOsys/grevaluator. API-level documentation can be found at https://ceosys.readthedocs.io/.

