## Supplementary Appendix for "Design and implementation of a system for automated monitoring of adherence to evidenced-based clinical guideline recommendations"

### Prototype Implementation

#### Implementation of the adherence evaluator

The adherence evaluator is the core module that reads the declarative machine-readable guideline recommendations and executes the rules defined therein on patient data to evaluate guideline recommendation applicability and adherence in individual patients. The adherence evaluator exposes a single API endpoint that triggers the retrieval of patient data and guideline recommendations and the evaluation of applicability and adherence. The results of these evaluations are provided to the user interface through a sub module, the user interface backend.

The logic of this core module is implemented in the Python package `cgr_adherence` (clinical guideline recommendation adherence; Figure 1). The package's main class `AdherenceEvaluator` is used to input guideline recommendations and executable rules from the recommendation as follows. First, two distinct collections of semantic terms are identified in the guideline recommendation: The semantic terms defining the population of patients to which the guideline recommendation is applicable (e.g., critically ill COVID-19 patients with need of oxygen supply), and the semantic terms defining the recommended intervention for these patients (e.g. daily administration of 1 mg Dexamethasone for 10 days). Then, for each collection of semantic terms, it is determined whether these describe a simple rule defined by a single entity (e.g., blood oxygen saturation below 90%) or whether the semantic terms describe a complex rule (e.g., a medication that is defined by multiple characteristics like the dose to be administered and the schedule, route and duration of administration). To enable the core module to apply the rules to the patient data, all semantic terms used in the machine-readable guideline recommendations are required to be coded using concepts from internationally standardized medical terminologies such as *Systematized Nomenclature of Medicine - Clinical Terms* (SNOMED CT) [1], and *Logical Observation Identifiers Names and Codes* (LOINC) [2], *International Statistical Classification of Diseases and Related Health Problems, 10$^{th}$ revision* (ICD-10) [3], *Anatomical Therapeutic Chemical Classification System* (ATC) [4] or *Unified Code for Units of Measure* (UCUM) [5] codes. To map these concept codes to the codes used in the clinical data, either a lookup table must be used (e.g., FHIR ConceptMap resources) or concepts codes from the guideline recommendation and the clinical data must use identical code systems (e.g. the OMOP Standard Vocabulary). Finally, `Quantity` objects are instantiated to represent the identified rules. These objects implement a `valid()` method that, when applied to patient data, indicates whether the rule represented by the `Quantity` object is followed in the data or not (e.g., whether the patient has a blood oxygen saturation <90%). In that way, the individual applicability and adherence of guideline recommendations can be checked most parsimoniously using a combination of `Quantity` objects representing the population (i.e., to which patients the recommendation is applicable to) and the intervention (i.e., which action to take or refrain from for these patients) part of the guideline recommendation, respectively.

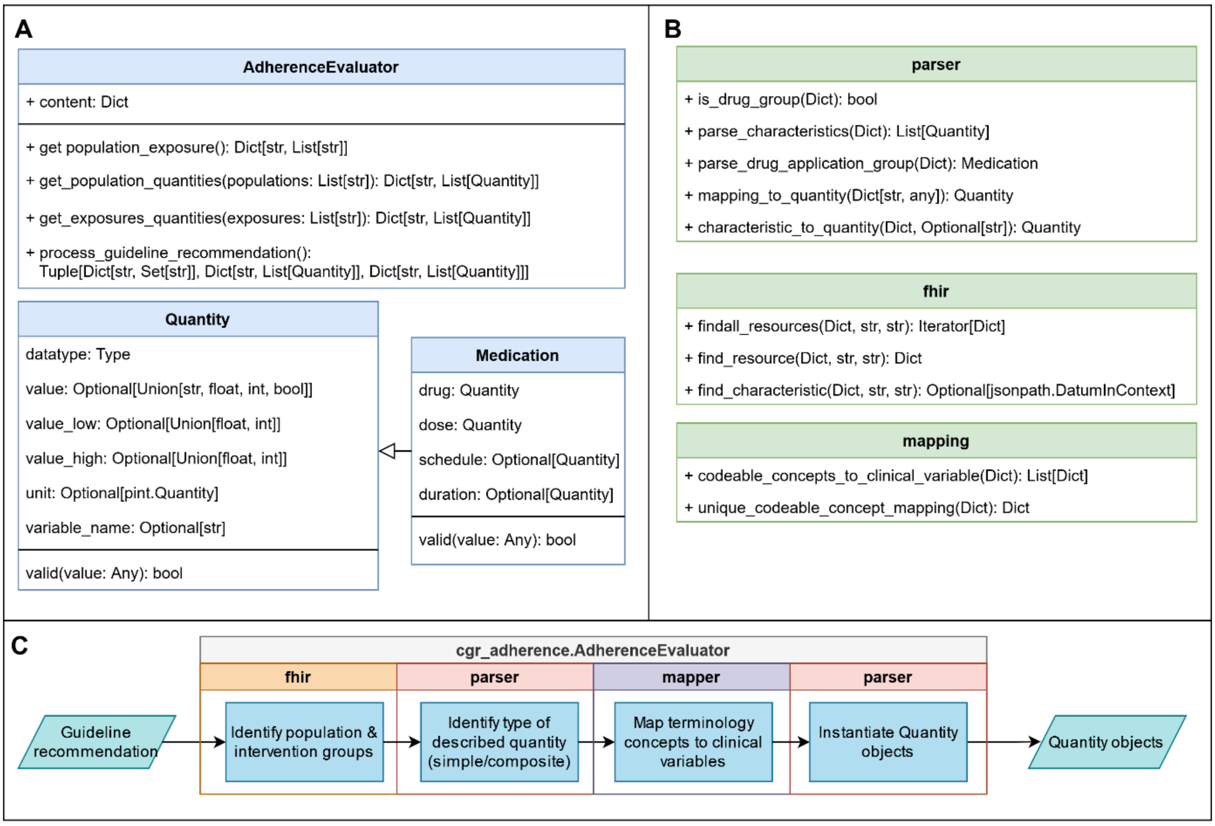


***Figure 1: Class diagram and flow chart of the clinical guideline recommendation adherence evaluator Python package****.* ***A*** *Shown are the main package class AdherenceEvaluator that performs the processing of guideline recommendations to output rules in the form of Quantity objects. Top cells depict public class fields, bottom cells public class methods.* ***B*** *The AdherenceEvaluator class uses accessory modules that perform the parsing of the guideline recommendation format and the mapping to clinical variables of the internal data format. Modules are shown with their public methods.* ***C*** *Shown is the process of the AdherenceEvaluator class from input of a guideline recommendation to output of guideline recommendation rules in the form of* `Quantity` *objects.*

The adherence evaluator module thus translates guideline recommendations into a ruleset in the form of a collection of `Quantity` objects. The module then requests all clinical data variables required by the `Quantity` objects to validate the rule it represents from the clinical data interface. It then applies the groups of population `Quantity` objects and intervention `Quantity` objects to the data returned by the clinical data interface to evaluate individual applicability and adherence of the guideline recommendation, respectively, using the `Quantity.valid()` methods. The individual results of this evaluation are then stored to be served by the user interface backend to the user interface.

The user interface backend provides external access to the results of the adherence evaluator through a RESTful API for the user interface to display and interact with these results. As this module provides the interaction with the "outside world", user authentication and authorization is performed at this stage to prevent unauthorized access to patient data and/or evaluation results. Thus, the user interface backend exposes an API endpoint for acquiring an access token using OAuth2-based [6] authentication in addition to endpoints related to retrieving the results of the adherence evaluation and endpoints related to retrieving individual patient data to allow the user interface to visualize the original patient data on which the adherence evaluation was performed.

### Implementation of the clinical data interface

The clinical data interface provides access to patient data in a standardized format to downstream modules. It exposes three main API endpoints for retrieving a list of current patients, for retrieving specific clinical variables (e.g., blood pressure or medication) for all current patients and for retrieving specific clinical variables from a specific patient.

The main task of the clinical data interface is to connect to individual hospital EMR systems to retrieve patient data and convert and clean these data into a standardized format for processing by the adherence evaluator module. In our prototype implementation for use at Charité - Universitätsmedizin Berlin, we have implemented an ETL (extract, transform, load) connector for three different hospital EMR systems. This connector retrieves, merges and cleans data and then converts it into the standardized format used by the adherence evaluator. Here, data may also be provided in another standardized format such as the OMOP common data model (CDM), to be consumed by the adherence evaluator. For demonstration purposes, the module implementation in our public repository instead passes a pre-defined list of patient data. This mock implementation needs to be adapted to the characteristics of individual hospital systems for use with real-time clinical data.

### Implementation of the guideline interface

The guideline interface provides clinical guideline recommendations in a machine-readable, declarative format to downstream modules. It exposes two main API endpoints, one for retrieving a list of available guideline recommendations and the second one for retrieving a specific guideline recommendation.

For the machine-readable specification of clinical guideline recommendations, we have developed and applied a FHIR format based on Evidence-Based Medicine on Fast Healthcare Interoperability Resources (EBMonFHIR), which is revised regularly and integrated with current developments of the

FHIR standard [7,8]. As the FHIR-based machine-readable guideline recommendation format is extensible and universal, any clinical guideline recommendation can be specified in that format and it is thus not limited to specific clinical guideline recommendations.

We designed our prototype system to retrieve machine-readable guideline recommendations hosted on MAGICapp (https://app.magicapp.org), the core platform of the MAGIC ("Making Grade the Irresistible Choice") Evidence Ecosystem Foundation, an independent non-profit organization that promotes the generation of evidence summaries and clinical guideline recommendations [9]. However, any other suitable hosting platform could be integrated in implementations of alternative guideline interface modules instead, including local hosting of guideline recommendations within hospitals' networks.
